# COVID-19 Vaccine Acceptance and Hesitancy in Ghana: A Systematic Review

**DOI:** 10.1101/2023.09.19.23295768

**Authors:** Godwin Banafo Akrong, Rosemond Akpene Hiadzi, Antonia Bernadette Donkor, Daniel Kwasi Anafo

## Abstract

The propensity to accept vaccines and factors that affect vaccine acceptance and hesitancy will determine the overall success of the COVID-19 vaccination program. Therefore, it is essential for countries to understand the factors that influence vaccine acceptance and hesitancy in order to prevent further future shocks, and it is necessary to have a thorough understanding of these factors. This study, as a result, aims to review selected published works in the domain of study and conduct valuable analysis to determine the most influential factors in COVID-19 vaccine acceptance and hesitancy in Ghana. The review also explored the acceptance rate of COVID-19 vaccines in Ghana. We selected published works from 2021 to April 2023 and extracted, analyzed, and summarized the findings based on the key factors that influence COVID-19 vaccine acceptance and hesitancy in Ghana, the acceptance rate in Ghana, the demographic factors that are often examined, and the study approach used to examine these factors. The study found that positive vaccination perception, safety, belief in vaccine efficacy, knowledge of COVID-19, and a good vaccine attitude influence COVID-19 vaccine acceptance in Ghana. The negative side effects of the vaccines, mistrust in the vaccine, lack of confidence in the safety of the vaccines, fear, and spiritual and religious beliefs all played significant roles in the factors influencing COVID-19 vaccine hesitancy. The demographic parameters frequently included in these studies that have a significant impact include educational attainment, gender, religious affiliation, age, and marital status.

## 1. Introduction

The COVID-19 pandemic has emerged as a worldwide public health crisis, presenting substantial challenges to healthcare systems and governments [1]. This resulted in the implementation of individual and community measures, such as improved hand hygiene practices, adherence to physical distancing guidelines, and the utilization of personal protective equipment, to mitigate the spread of diseases.

In the wake of the COVID-19 pandemic, the Africa Center for Disease Control implemented the SARS-COV-2 Development and Access Strategy in 2020. The primary objective of this strategy is to achieve vaccination coverage of at least 60% among the African population by 2022 to attain herd immunity [2, 3]. According to the World Health Organization [4], a total of 53 out of the 54 African countries have initiated the COVID-19 immunization campaign. As of the latest data from the WHO [4], these countries have collectively received 1,114,404,185 cumulative doses. Out of this total, 851,588,735 doses, or approximately 76%, have been administered. According to the World Health Organization (WHO), the administration rate of vaccine doses in Africa stands at 61 per 100 people. Approximately 39.4% of the African population, or 552 780 583 individuals, have received at least one dose of the vaccine, according to the most recent data. Furthermore, 32.6% of the population, equivalent to 458,147,110 individuals, has completed the primary series of vaccinations. This suggests that there is a necessity to enhance vaccination coverage among Africans, particularly among individuals who have received only one dose and those who have not been vaccinated at all. Encouraging these individuals to finish their vaccination regimen would contribute to the attainment of the target established by the Africa Center for Disease Control in 2020.

This observation underscores the importance for African nations to prioritize the study of factors influencing individuals’ acceptance or hesitancy towards COVID-19 vaccines. Understanding these factors is crucial, as they are believed to impact both those who have received only one dose and those who have not yet been vaccinated. It is important to acknowledge that Ghana, situated in West Africa, is actively engaged in this endeavor. Based on the World Health Organization’s report in 2023, it is evident that all 17 West-African countries have initiated the COVID-19 immunization campaign [4]. These countries have collectively received a total of around 337,393,634 vaccine doses, out of which 257,991,442 doses (76%) have been administered. Notably, the vaccination rate in West Africa is 55 doses per 100 people. It is worth noting that a significant proportion, specifically 36.8%, of the countries situated in the West African region have been administered a minimum of one dose of the COVID-19 vaccine. Furthermore, around 30.8% of the population, which amounts to 143,800,552 individuals, has completed the initial series of vaccinations. Nevertheless, it is imperative to implement additional steps to ensure that those who have received only one dose of the vaccine follow through with completing the whole vaccination regimen. Moreover, it is crucial to establish mechanisms that encourage individuals who have not yet received any doses of the vaccine to do so. The primary objective of this study is to carry out an in-depth review, analysis, and assessment of the acceptability and hesitancy of the COVID-19 vaccine in Ghana. This is to assist in determining the factors that influence COVID-19 vaccine acceptance and COVID-19 vaccine hesitancy, with a focus on Ghana.

Per the World Health Organization [4], Ghana has received a total of 34,047,598 cumulative doses of vaccines. Out of these doses, they have successfully administered 25,624,828, which accounts for almost 75% of the total received. Consequently, there is a remaining quantity of around 8,422,770 doses yet to be administered. Nevertheless, the data obtained from the World Health Organization (WHO) provides additional evidence that a total of 13,864,186 individuals, accounting for approximately 43.7% of the population in Ghana, have gotten at least a single dose of the vaccine. This indicates that a greater number of vaccines will be required for individuals to achieve full COVID-19 vaccination, despite 10,780,003 individuals having already finished the vaccination process. One notable aspect of Ghana is the high rate of vaccine administration, with 81 doses being delivered per 100 individuals in the population. However, the objective is to achieve widespread acceptance of the COVID-19 vaccination throughout the general population while also identifying the variables that hinder their acceptance of it. This study aims to conduct a systematic review to determine the prevailing factors that researchers have identified as significant contributors to COVID-19 vaccine acceptance in Ghana, as well as the factors that contribute to vaccine hesitancy among the Ghanaian population. This can serve as a foundation for motivating individuals who have not yet received the vaccine or who have not yet received their second dose.

A study conducted by Amponsah-Tabi et al. [5] during the period of the COVID-19 vaccine rollout examined the influence of media awareness on the acceptance of vaccination. This study highlighted that those who had prior knowledge or education about vaccination were influenced by the inclusion of media awareness, hence influencing the dynamics of COVID-19 vaccine acceptability. The results of the study provided more evidence that the acceptance, understanding, and perception of COVID-19 vaccination remain limited among individuals residing in rural communities in Ghana. Therefore, it was imperative to provide education to individuals residing in remote villages regarding the advantages of the COVID-19 vaccine to attain a successful vaccination initiative. Additionally, the study recognized that individuals’ prior vaccination history played a significant effect on their willingness to accept the COVID-19 vaccine. Consequently, it is recommended that national vaccination initiatives, such as the Hepatitis B program, be utilized as a means to enhance the acceptance of the COVID-19 vaccine. Adjaottor et al. [6] indicated that parents and guardians should take a bigger role in helping their children make the right COVID-19 vaccination decisions while assessing the acceptance of the vaccine among teenagers in Ghana. Additionally, it is worth noting that the majority of studies investigating the factors influencing the acceptability of COVID-19 vaccines in Ghana were completed before the commencement of the vaccination campaign [7]. The study additionally asserts that vaccine acceptance decisions exhibit temporal variation, suggesting that individuals’ decisions may have changed after the availability of vaccines. Therefore, the Government of Ghana must adopt measures aimed at promoting favorable attitudes towards vaccinations, heightening the perception of danger associated with getting the virus, and educating the general public about the advantages of vaccination.

Healthcare professionals have been identified as a high-risk population for contracting and experiencing mortality due to COVID-19 [8]. Therefore, the crucial aspect lies in the acceptance and use of the COVID-19 vaccination, among healthcare professionals as it plays a pivotal role in safeguarding their well-being and safety. The results of their study further demonstrated that younger healthcare personnel, non-Christians, and those employed in facilities with a religious affiliation were more inclined to take part in a COVID-19 vaccine acceptance when given the chance. The study conducted by Forkuo et al. [9] also underscores the importance of possessing sufficient knowledge about COVID-19 and vaccines as a key factor in promoting vaccine acceptability in Ghana. It further indicates that campaign messages designed to enhance COVID- 19 vaccine coverage should prioritize the communication of its safety, potential side effects, and proper management. Addressing the misconceptions surrounding these factors will contribute to the eradication of false beliefs among the population.

Additional research findings indicate that the health management team has the potential to mitigate vaccine hesitancy by prioritizing efforts to minimize the adverse influence of the media and other factors such as fear and mistrust in leadership [10]. Furthermore, it has been observed that older age groups in Ghana have low levels of vaccine hesitancy although males tend to be more indecisive about receiving the vaccine compared to females [11]. Nevertheless, the study did not uncover any noteworthy correlations between the inclination to vaccinate and factors such as level of education or geographical location.

Alhassan et al. [12] acknowledged that fear; uncertainty, conspiracy theories, and safety concerns pose significant challenges to the effective adoption of the vaccine if not well addressed. Therefore, according to Botwe et al. [13], safety education must also make an effort to allay the fears of those who have had negative reactions to other vaccines, those who believe the vaccination is intended to make people infertile, and those who have underlying medical conditions but were unsure they could take the vaccines. According to Amo-Adjei et al. [14], respondents’ socioeconomic characteristics were less predictive of acceptance than their confidence in the vaccine’s efficacy and safety. The authors additionally assert that a lack of faith in domestic and international political figures, a belief in divine protection, and a perceived lack of awareness regarding the processes involved in vaccine production all contribute to the rejection of vaccines. Additional research conducted by Mohammed et al. [15] indicates that several variables are significant indicators of individuals’ likelihood of receiving the COVID-19 vaccine. These factors include being between the ages of 25 and 45, being over the age of 45, identifying as male, and identifying as Christian. However, several individuals expressed concerns about not receiving vaccines, citing factors such as the perceived quick development and approval process, potential immediate side effects, and unanticipated long-term consequences.

Despite the existence of numerous surveys and literature examining the factors that influence the acceptance and hesitancy of COVID-19 vaccines, there is a lack of comprehensive systematic reviews in the field of vaccine acceptance that have endeavored to uncover the factors influencing both acceptance and hesitancy of COVID-19 vaccines. Based on Kitchenham et al. [16] evaluation checklist, it was evident that a sizable portion of the researchers who carried out these investigations in Ghana primarily presented a narrative review rather than a systematic review. Their reviews encompassed broad insights into the differing determinants of COVID-19 vaccine acceptance and the factors contributing to COVID-19 hesitancy. Furthermore, it is important to highlight that there is currently no available systematic review that examines the various factors that impact the acceptance or hesitancy of the COVID-19 vaccine in one paper. This knowledge gap serves as motivation for our study, as previous studies have aimed to offer a comprehensive review, analysis, and evaluation of the patterns of COVID-19 vaccination uptake and hesitancy specifically within the context of Ghana. Multiple systematic studies have been conducted to comprehensively examine global data on COVID-19 epidemics and the determinants of national COVID-19 vaccination rates [17–20]. The decision to conduct a systematic review was influenced by the existing knowledge in the field. This review seeks to determine the factors influencing COVID-19 acceptance and hesitancy in Ghana, as well as the demographic characteristics that are commonly assessed in relation to COVID-19 acceptance and hesitancy. Additionally, the review will examine the prevailing research methodologies employed in this area of study. Furthermore, the review will provide an analysis of the COVID- 19 vaccine acceptance rate in Ghana from 2021 to the first quarter of 2023.

The subsequent sections of this paper are organized in the following manner. Section 2 provides a comprehensive account of the methodology employed in the present review. The findings and analysis are presented in Section 3, while the implications and potential areas for further research are discussed in Section 4.

## 2. Methodology

The primary aim of this study is to provide a comprehensive review, analysis, and evaluation of the COVID-19 vaccination uptake and hesitancy in Ghana. The present study offers a contemporary overview and evaluation of several key aspects: (1) the factors that contribute to COVID-19 vaccine acceptance in Ghana; (2) the factors that contribute to COVID-19 vaccine hesitancy; (3) the demographic profile that is most frequently assessed and its association with acceptance and hesitancy of COVID-19 vaccine in Ghana; (4) the predominant study methodology employed in this field; and (5) the acceptance rate of COVID-19 in Ghana. Table 1 presents the five research questions that have been formulated to conduct this systematic literature review. This systematic review was carried out following the guidelines outlined by Preferred Reporting Items for Systematic Review and Meta-Analysis (PRISMA) [21]. The review encompassed the identification, screening, and inclusion stages as per the recommended protocols.

**Table 1.** Systematic review questions.

| # | Question | Solution |
| --- | --- | --- |
| RQ1 | What are the factors that contribute to the acceptance of the COVID-19 vaccine in Ghana? | Identify the factors that contribute to the COVID-19 vaccine acceptance in Ghana. |
| RQ2 | What are the contributing causes of COVID-19 vaccine hesitancy in Ghana? | Explore the factors that contribute to Ghana's COVID-19 vaccination hesitancy. |
| RQ3 | Which demographic factors are commonly evaluated while determining the factors that influence the acceptance and hesitancy of the COVID-19 vaccine in Ghana? | Review the demographic factors that are typically considered when determining the factors that influence the acceptance and hesitancy of the COVID-19 vaccine in Ghana. |
| RQ4 | What are the main study approaches utilized in examining the factors influencing the acceptance and hesitancy of the COVID-19 vaccine in Ghana? | Determine the most prevalent study methodology utilized by researchers in determining the factors that affect the acceptance and hesitancy of COVID-19 in Ghana. |
| RQ5 | What is the COVID-19 vaccine acceptance rate in Ghana? | Examine the rate of COVID-19 vaccine acceptance in Ghana. |

Initially, we developed specific research questions that align with the objective of this study. The next step is to develop the search strategy, which entails choosing the search phrases and appropriate search engines that can be used to get relevant literature for an additional search. To identify the relevant studies that contribute to solving the research inquiries, a criterion for study selection was established. In order to refine the criteria for study selection, a preliminary pilot study selection was conducted. Subsequently, some quality checklists were implemented to evaluate relevant studies as part of the quality evaluation procedure. The data that was gathered was synthesized. The selection of suitable synthesis approaches was guided by the nature of the data and the research questions that were addressed through the data collection process. Figure 1 illustrates the review protocol.

**Figure 1.**
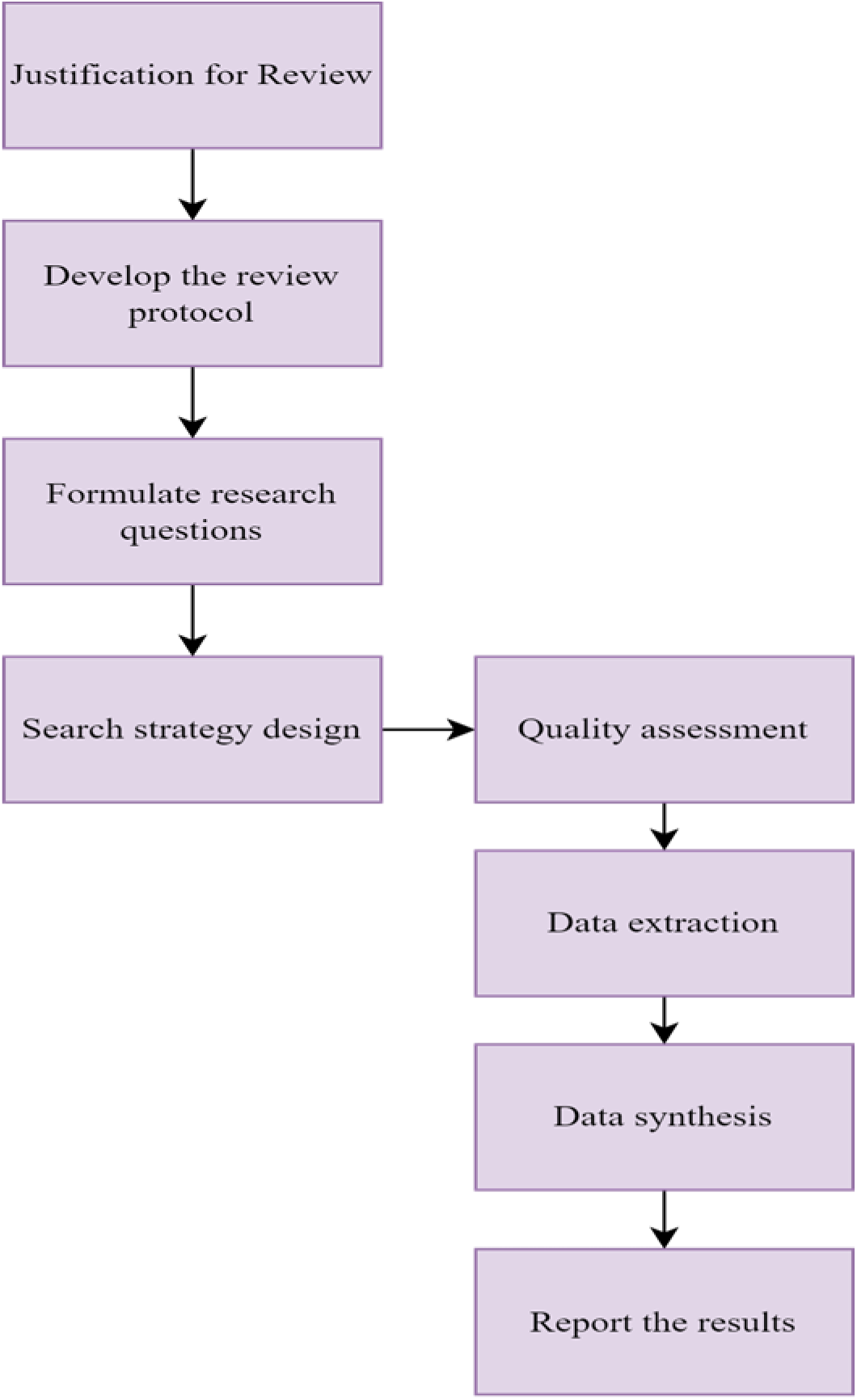
The review protocol.

### 2.1 Search Strategy

The components of the search strategy, namely the search phrases, search engines, and search strategy, will be elaborated upon in the subsequent sections.

#### 2.1.1 Search Terms

The search phrases included in this study were determined following the methodology outlined by Kitchenham et al. [16].

i. The process of identifying relevant keywords from scholarly papers.
ii. Employing alternate synonyms for the identified keywords.
iii. Generating key terms based on the research questions.

The search method employed in this study was the utilization of specific keywords and phrases such as “vaccine acceptance,” “novel coronavirus,” “vaccines,” “vaccination,” “COVID vaccine,” “vaccine hesitancy,” “COVID-19,” “COVID vaccine acceptance,” “Ghana,” and “level of COVID-19 vaccine acceptance and vaccine hesitancy.” The search string utilized for querying search engines followed a specific procedure, consisting of the following terms: (i) COVID AND vaccine AND acceptance, (ii) COVID AND vaccine AND hesitancy, (iii) level AND of AND COVID-19 AND vaccines AND acceptance, and (iv) novel AND COVID-19 AND hesitancy. The search string that was developed was designed to strike a balance between being of a manageable size and providing sufficient coverage.

#### 2.1.2 Search Engines

Following the establishment of the search terms, the selection process was undertaken to identify suitable and relevant search engines. The choice of search engines was not limited based on their accessibility at the researcher’s home institution. The process of identifying primary studies was conducted by searching several databases, including PUBMED, SpringerDirect, ProQuest, Medline, WorldCat Discovery, Embase, EBSCOhost, and ScienceDirect. The search strings generated were utilized to conduct searches in the aforementioned databases, specifically targeting articles. The search was further refined to encompass the time frame from January 1, 2020, to April 2023, inclusively. This temporal restriction was implemented to explore the most recent advancements and patterns pertaining to the acceptance and hesitancy surrounding COVID-19 in Ghana.

#### 2.1.3 Search Process

A first exploratory search was undertaken to assess the adequacy of available literature resources for the study area at hand. Once the search engines and search strings had been identified and defined, a thorough search was conducted on each of the eight electronic databases individually to retrieve relevant articles. The literature resources that were obtained by the search were downloaded and afterward exported into Excel. In the case of ScienceDirect, the software program JabRef (https://www.jabref.org/) was employed as an alternative to Excel. This choice was made due to the absence of an export-to-Excel feature in the electronic database, as well as the limitation of being able to download and export just 100 articles at a time. In the long run, the articles that were downloaded were subsequently combined and exported to Excel to conduct a human review and selection process to identify the relevant articles. The software package Mendeley (https://www.mendeley.com/) was employed to save and manage relevant articles. Furthermore, the search parameters were restricted to exclusively English-language articles, with the geographical scope confined to retrieving materials solely from Ghana.

### 2.2 Study Selection

The study selection process excluded article papers that do not offer relevant information for addressing the research questions outlined in this review. The selection process consisted of two parts. In the first phase, known as selection stage 1, papers were assessed to determine their relevance in addressing the study questions. Any articles deemed irrelevant were deleted from consideration based on predetermined inclusion and exclusion criteria. The second stage of the selection process involved the utilization of selection criteria to identify papers that were deemed relevant and of high quality. A total of 784 articles were generated as a result of the search process. The articles underwent a process of selection and elimination, which involved scrutinizing the titles, removing any duplicates, selecting potentially relevant papers based on abstract scrutiny and inclusion criteria, and conducting a thorough review of the selected papers for quality assessment. The review approach involved the selection of relevant research papers of satisfactory quality, which were subsequently utilized for data extraction. To identify appropriate papers for inclusion in the systematic review, the search criteria were implemented based on the specified research questions. Figure 2 depicts the search methodology and the progressive count of publications identified at each stage.

**Figure 2.**
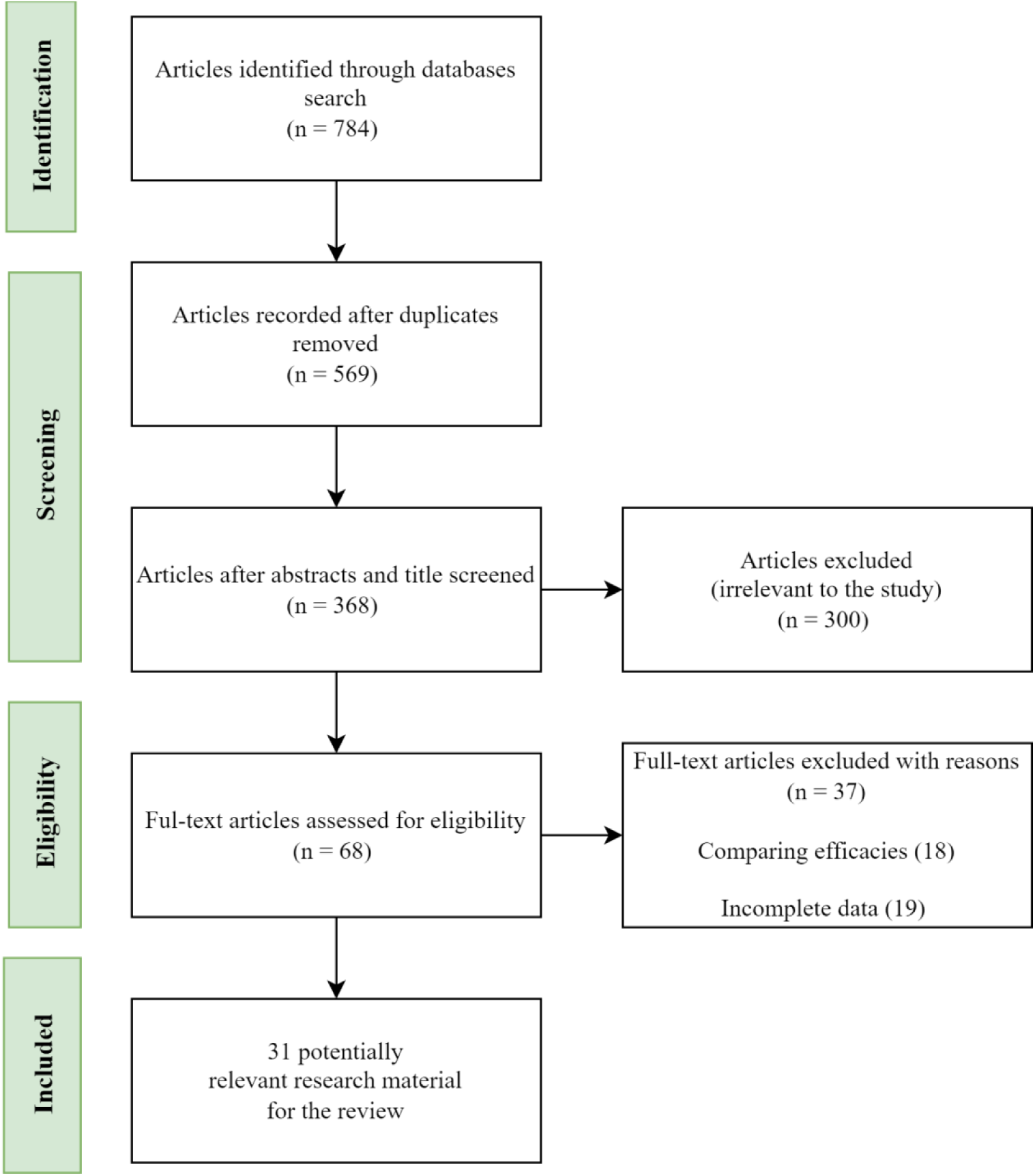
Flow diagram of the article selection process.

#### 2.2.1 Inclusion Criteria

The procedure includes the inclusion of articles sourced from academic journals, including but not limited to the following:

i. Articles with the primary goal of examining the factors that contributes to COVID-19 vaccine acceptance and hesitancy.
ii. Articles published between 2021 to April 2023 were included.
iii. Articles used in the study should focus on Ghana.
iv. Papers written in English were included.

#### 2.2.2 Exclusion Criteria

Excluded from consideration were papers that consisted just of an extended abstract or a PowerPoint presentation.

i. Books and magazines were omitted.
ii. In addition, review papers were omitted from consideration, resulting in a total of 68 articles that were potentially relevant after undergoing a quality assessment evaluation. Following the evaluation of eligibility based on the 68 articles, a total of 31 final relevant studies were identified.

### 2.3 Study Quality Assessment

During the evaluation process, measures were implemented to ensure the quality of the search. The review of articles was conducted manually following the initial automatic search. Subsequently, we conducted a thorough examination and analysis of the titles and abstracts to ascertain the suitability of the papers for inclusion or exclusion. To mitigate the potential impact of prior search history, the initial search for articles was performed in private browsing mode. Furthermore, a comprehensive questionnaire was developed to evaluate the relevance of the studies that were incorporated. The purpose of formulating these questions was to assess the pertinence, thoroughness, and reliability of the papers. Several of these questions were formulated using the research conducted by Wells and Littell [22]. Each question was assigned a score based on one of three alternative answers: “yes” with a score of 1, “partly yes” with a score of 0.5, and “no” with a score of 0. The cumulative scores were obtained by aggregating the scores derived from responses to the quality evaluation questions, representing the quality score assigned to each of the studies included in the analysis. Only papers with a quality score of 5 or higher were included in the data extraction and synthesis processes.

### 2.4 Data Extraction

A data form was created in Microsoft Excel to systematically gather data that addresses the review questions outlined in each included paper. This was accomplished by utilizing Table 2. We have compiled a summary of data on COVID-19 vaccine acceptance, COVID-19 vaccine hesitancy, acceptance rates, study duration, and geographical location. The collection process also included gathering standard information such as publication details, publication date, title, author name or names, and publication location. During the process of extracting data, it was noted that not all of the studies included in the analysis were able to address all of the research questions posed in the review. An additional challenge discovered throughout the process of extracting data was the presence of varying terminologies utilized across different studies. In certain academic works, the terms “novel covid19” and “SARS-CoV-2” have occasionally been employed as substitutes for the term “COVID-19.” In order to mitigate any problems related to ambiguity, we have chosen to utilize the term “COVID-19” for all.

**Table 2.**
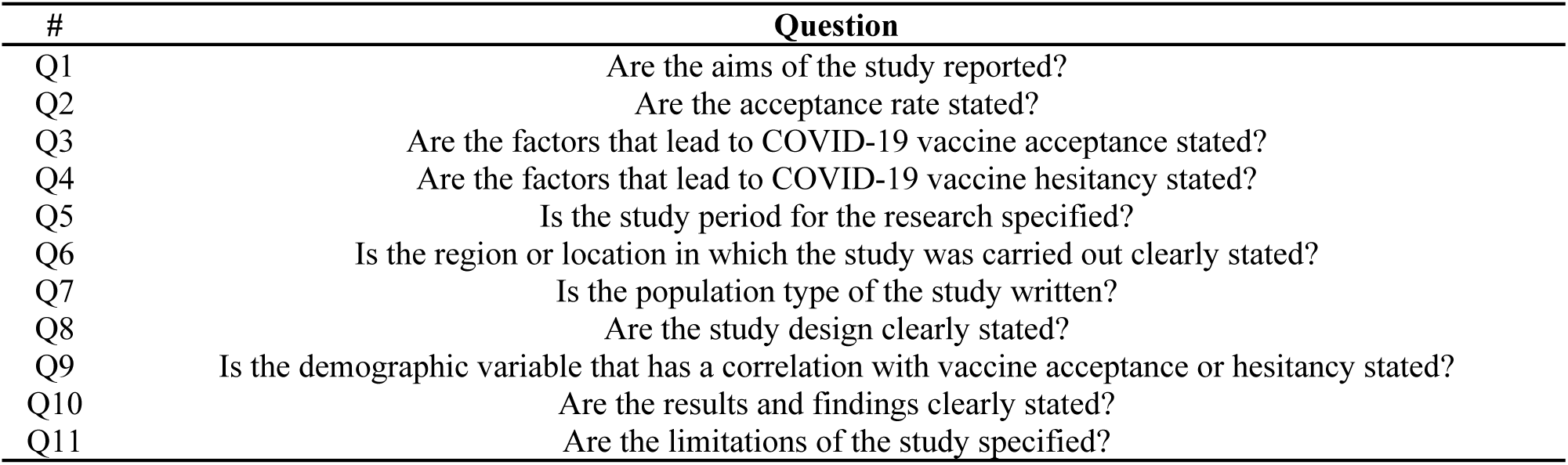
Quality assessment questions.

### 2.5 Data Synthesis

The data that was gathered was saved for utilization in the step of synthesizing the data. The primary objective of data synthesis is to consolidate and summarize the gathered data from the included studies to offer comprehensive responses to the established review questions. The findings from all the included studies that offer comparable or equivalent evidence were added up to arrive at a conclusion. The review involved the extraction of quantitative data, which were subsequently synthesized to present our findings in a comparable manner [23]. Furthermore, we employed a narrative synthesis approach in response to the data obtained from our review questions. Consequently, a variety of visualization approaches were employed, including doughnut charts, clustered bar graphs, line graphs, clustered columns, and pie charts. Finally, tables were utilized to summarize and illustrate the findings.

## 3. Results and Discussions

### 3.1 Description of the Included Studies

This section provides a concise summary of the studies that have been included. A total of 31 scholarly articles pertaining to the subject of COVID-19 vaccine acceptance and vaccine hesitancy were identified, encompassing the time frame from 2021 to April 2023, inclusive. Table 3 lists the research questions that each of the included papers addressed.

**Table 3.**
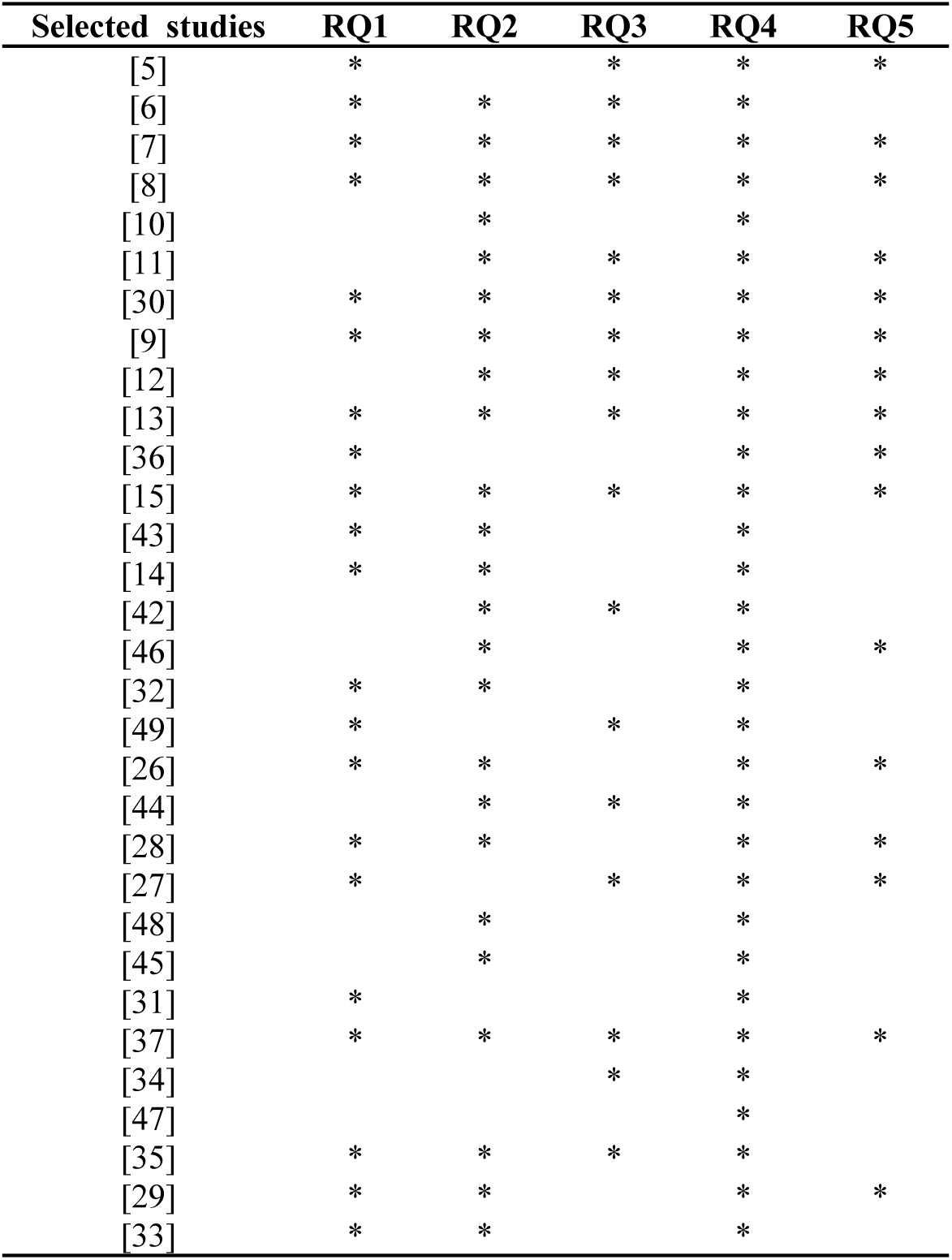
Research questions answered in included studies.

| Selected studies | RQ1 | RQ2 | RQ3 | RQ4 | RQ5 |
| --- | --- | --- | --- | --- | --- |
| [5] | * |  | * | * | * |
| [6] | * | * | * | * |  |
| [7] | * | * | * | * | * |
| [8] | * | * | * | * | * |
| [10] |  | * |  | * |  |
| [11] |  | * | * | * | * |
| [30] | * | * | * | * | * |
| [9] | * | * | * | * | * |
| [12] |  | * | * | * | * |
| [13] | * | * | * | * | * |
| [36] | * |  |  | * | * |
| [15] | * | * | * | * | * |
| [43] | * | * |  | * |  |
| [14] | * | * |  | * |  |
| [42] |  | * | * | * |  |
| [46] |  | * |  | * | * |
| [32] | * | * |  | * |  |
| [49] | * |  | * | * |  |
| [26] | * | * |  | * | * |
| [44] |  | * | * | * |  |
| [28] | * | * |  | * | * |
| [27] | * |  | * | * | * |
| [48] |  | * |  | * |  |
| [45] |  | * |  | * |  |
| [31] | * |  |  | * |  |
| [37] | * | * | * | * | * |
| [34] |  |  | * | * |  |
| [47] |  |  |  | * |  |
| [35] | * | * | * | * |  |
| [29] | * | * |  | * | * |
| [33] | * | * |  | * |  |

### 3.2 Publication Year

Figure 3 shows the distribution of articles published between the years 2021 to April 2023. In general, the distribution exhibits a marginal decrease in the research trajectory within the field of study. The doughnut charts illustrate a progressive rise in the publication of research about the determinants of COVID-19 vaccine uptake and COVID-19 vaccination hesitancy spanning the period from 2021 to 2022. Between the periods of 2022 to April 2023, a little decline has been observed in the number of articles gathered about Ghana. The decline observed in the current season may be attributed to the progressive interventions implemented to combat the COVID-19 pandemic and promote vaccine acceptance within the Ghanaian population. The study posits that there exists a proclivity for an increased number of research articles to be conducted before the end of the year 2023.

**Figure 3.**
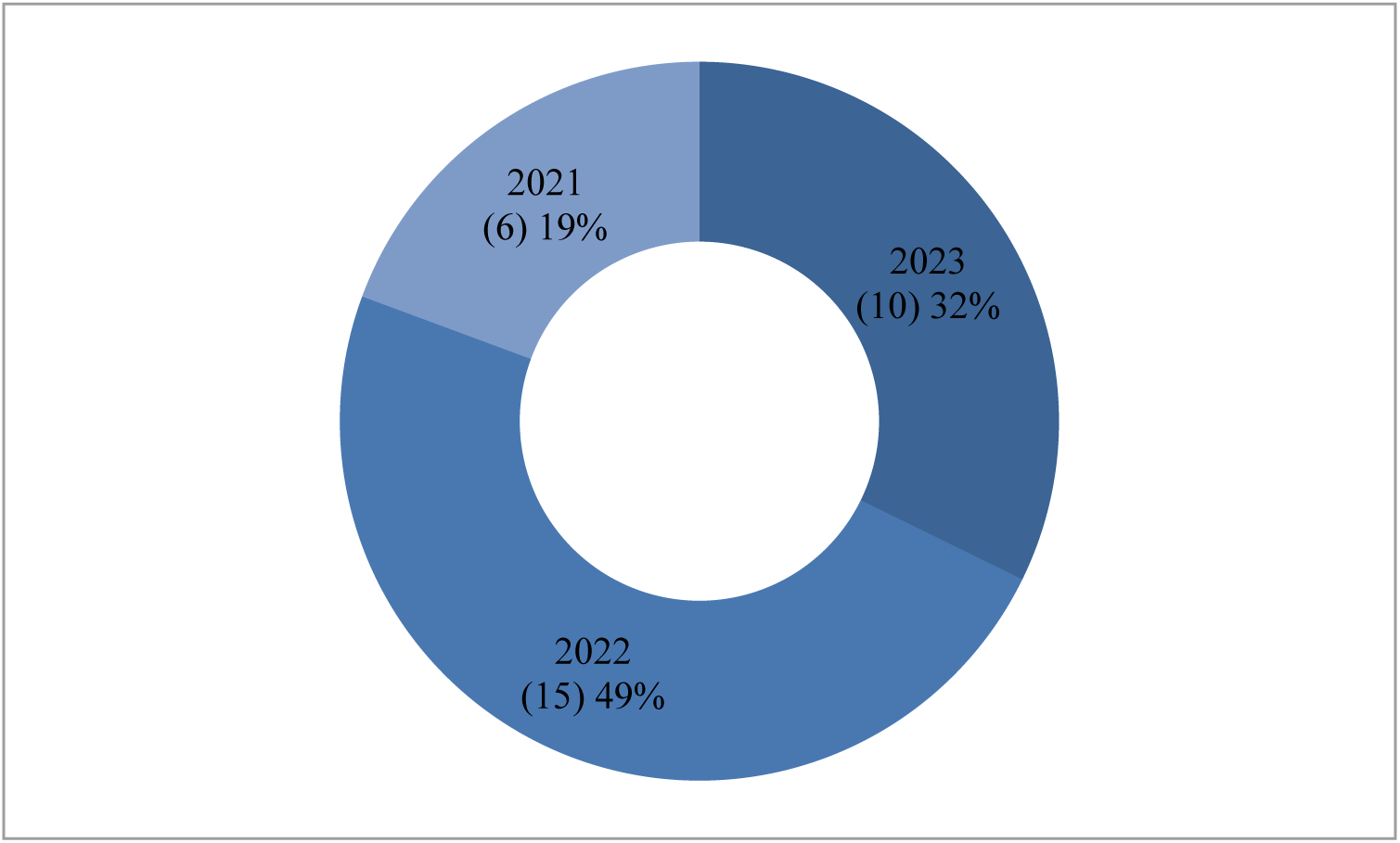
Publication trend from the year 2021- April 2023

### 3.3. Publication Source

Table 4 provides a summary of the articles, including the number of primary studies, in the respective journal. The studies incorporated in this analysis were published in a total of nineteen distinct academic journals. PloS One stands out as the journal with the highest number of publications, totaling five. Subsequently, the aforementioned publications included Vaccines (3), Advances in Public Health (3), and Ghana Medical Journal (3).

**Table 4.** Summary of journal publications (2021– April 2023).

| **Journal name** | **Frequency** |
| --- | --- |
| PLoS One | 5 |
| Vaccines | 3 |
| Advances in Public Health | 3 |
| Ghana Medical Journal | 3 |
| International Journal of Environmental Research and Public Health | 2 |
| Archives of Public Health | 2 |
| BMC Infectious Diseases | 1 |
| Human resources for health | 1 |
| Tropical Medicine and Health | 1 |
| Radiography | 1 |
| Clinical and Experimental Vaccine Research | 1 |
| Human Vaccines & Immunotherapeutics | 1 |
| Health Science Reports | 1 |
| The Pan African Medical Journal | 1 |
| Public Health Challenges | 1 |
| Health Research Policy and Systems | 1 |
| Therapeutic Advances in Vaccines and Immunotherapy | 1 |
| Risk Management and Healthcare Policy | 1 |
| African Geographical Review | 1 |

### 3.4 Analysis on Systematic Review Questions

#### 3.4.1 What are the factors that contribute to the acceptance of the COVID-19 vaccine in Ghana?

Based on the reviewed literature, Table 5 outlines the factors that contribute to the acceptance of the COVID-19 vaccine in Ghana. The findings show that the acceptance of the COVID-19 vaccine in Ghana is primarily attributable to the citizens’ positive perception of the vaccine, the vaccine’s safety, and their confidence in its efficacy. In addition, it emerged that knowledge of COVID-19 and a positive attitude toward the vaccine play a significant role in the acceptance of the COVID-19 vaccine in Ghana. Several additional factors were found to be associated with the acceptance of the COVID-19 vaccine. These factors include the desire to safeguard oneself and one’s family, the gender of the participants, the influence of observing others receiving the vaccine, the respondents’ willingness to be vaccinated, and their perception of susceptibility to the virus, marital status, religious affiliation (Christianity), and their willingness to pay for the vaccine. It was observed that while factors such as individuals’ highest level of education, employment as salaried workers, high levels of trust in the government, and recent receipt of vaccines such as HBV were also influential in the acceptability of the COVID-19 vaccine in Ghana, their impact was relatively small.

**Table 5.** Reasons for accepting COVID-19 vaccine.

| # | Reasons | Studies | Frequency |
| --- | --- | --- | --- |
| 1 | Good perception of vaccine | [5], [26-28] | 4 |
| 2 | Safety of the vaccine | [6], [13], [14], [29] | 4 |
| 3 | Trust in the effectiveness | [14], [27], [30], [31] | 4 |
| 4 | Knowledge of COVID-19 | [9], [32], [33] | 3 |
| 5 | Good attitude towards vaccine | [5-7], [27] | 3 |
| 6 | Protect oneself and family | [9], [28] | 2 |
| 7 | Gender | [15], [32] | 2 |
| 8 | Seeing others get the vaccine | [26], [29] | 2 |
| 9 | Willingness | [29], [34] | 2 |
| 10 | Believing COVID-19 information | [29], [35] | 2 |
| 11 | Perceived susceptibility | [7], [9] | 2 |
| 12 | Relative being diagnosed with COVID-19 | [29], [30] | 2 |
| 13 | Being married | [33], [36] | 2 |
| 14 | Receive other forms of vaccination twice | [26], [28] | 2 |
| 15 | Being a Christian | [8], [15] | 2 |
| 16 | Willing to pay for the vaccine | [9], [31] | 2 |
| 17 | Perceived benefit | [7] | 1 |
| 18 | Non-Christian | [8] | 1 |
| 19 | Vaccine being cost-free | [32] | 1 |
| 20 | Highest educational level | [33] | 1 |
| 21 | Receiving recent vaccines such as HBV | [5] | 1 |
| 22 | Working in faith-based health facilities | [8] | 1 |
| 23 | Trust in the accuracy of measures taken by government in the fight against COVID-19 | [37] | 1 |
| 24 | Salary workers | [36] | 1 |
| 25 | Test positive for COVID-19 | [26] | 1 |
| 26 | High trust in Government | [26] | 1 |

The aforementioned findings are consistent with the research by Hawlader et al. [24](2022), which established that people from Bangladesh, India, Pakistan, and Nepal demonstrated a notable tendency towards accepting vaccines due to their positive perception of vaccines and their concerns regarding potential side effects. Furthermore, it was observed that factors such as sex, marital status, education level, concerns about contracting the virus, perceived influence of COVID-19, beliefs regarding the effectiveness of vaccines, positive attitudes towards mandatory measures, and the availability of vaccines were significantly correlated with the acceptance of vaccines across various countries. The study conducted by Lee et al. [25](2022) provides additional support for the factors that have been recognized as significant in influencing the level of acceptance of the COVID-19 vaccines within the Ghanaian population. The evidence indicates that the level of intention to accept the vaccine was most pronounced among individuals who identified as female.

#### 3.4.2 What are the contributing causes of COVID-19 vaccine hesitancy in Ghana?

This section provides a summary of the multiple contributing factors to COVID-19 vaccine hesitancy in Ghana throughout the period spanning from 2021 to April 2023. The collected data indicates that the primary factor leading to COVID-19 vaccine hesitancy in Ghana is the presence of adverse side effects associated with the vaccines. Subsequently, a lack of trust in the vaccine and a lack of confidence in the safety of the vaccine, accompanied by fear and spiritual and religious convictions, as well as logistical inadequacies, emerged as contributing factors to COVID-19 vaccine hesitancy. Table 6 presents additional evidence indicating that a lack of sufficient knowledge about COVID-19 among individuals in Ghana, coupled with uncertainties and concerns related to conspiracy theories surrounding the impact of vaccines on the Ghanaian race, doubts regarding the efficacy of the vaccine, concerns about fertility implications, a lack of trust in state institutions, the perception of rapid development and approval of vaccines, prolonged waiting times at vaccination centers, gender disparities, educational attainment, safety concerns, traumatic stress, media attention, and political associations, collectively contributed to vaccine hesitancy in Ghana in relation to COVID-19. Nevertheless, it was found that factors such as residing in less urbanized areas, the politicization of response measures, marital status, disbelief in the existence of COVID-19, perceptions of low susceptibility to the virus, a preference to wait until the vaccine is perceived as safe, lack of trust in vaccines, and reliance on information from newspapers were also identified as contributing factors to vaccine hesitancy in Ghana. However, these factors were found to have a lesser impact compared to the earlier identified factors.

**Table 6.**
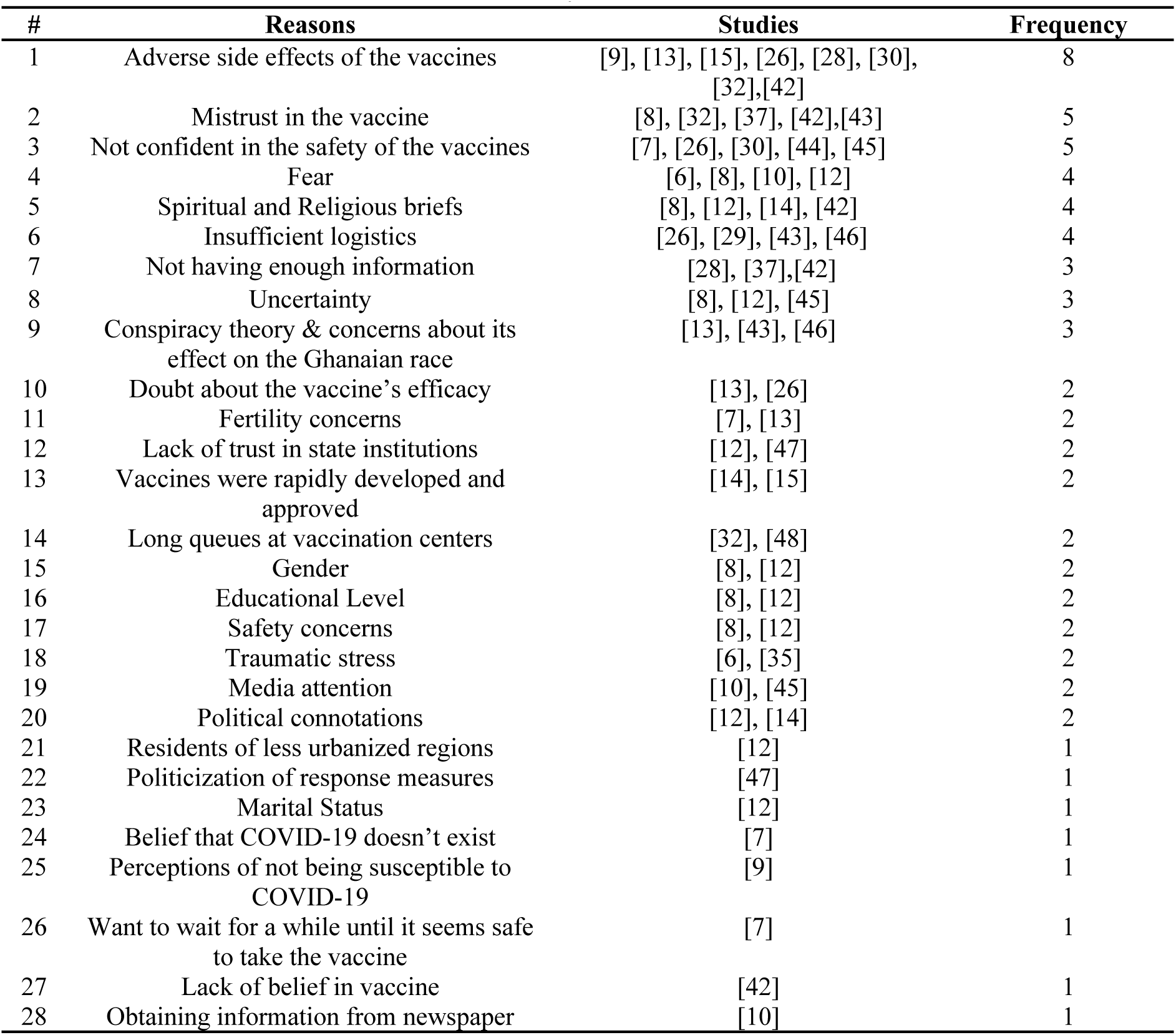
Reasons for COVID-19 vaccine hesitancy.

The contributing factors associated with vaccination hesitancy in relation to COVID-19, as identified in the conducted review, align with the findings of Diaz et al. [38](2022). The aforementioned study highlights that individuals who had not received the vaccine expressed concerns regarding potential long-term harmful effects that are currently unknown. Given their perspective, these individuals held the belief that COVID-19 vaccines may have adverse effects on reproductive health and fertility and expressed uncertainty regarding their potential influence on fertility. According to Freeman et al. [39](2022), there is evidence suggesting that hesitancy is more prevalent among those who identify as female, have lower income levels, and belong to certain ethnic groups.

#### 3.4.3 Which demographic factors are commonly evaluated while determining the factors that influence the acceptance and hesitation of COVID-19 vaccine in Ghana?

The significance of demographic analysis is in the provision of valuable information that can inform effective decision-making processes undertaken by governmental bodies and social service organizations [40, 41]. Additionally, it aids individuals in comprehending the attributes of a certain population and its potential future transformations, a crucial aspect in informing decision-making processes. The findings derived from the conducted review indicate that the demographic factors frequently examined in the assessment of the factors influencing the acceptance and hesitancy of COVID-19 in Ghana primarily revolve around educational attainment, gender, religious affiliation, age, marital status, and the primary source of information regarding the COVID-19 vaccine (see Table 7). It is crucial to recognize that demographic factors such as employment status, rural or urban residency, occupation, level of experience, and political party affiliation among opposition voters were not extensively examined in the assessment of factors influencing COVID-19 vaccine acceptance and hesitancy in Ghana (see Figure 4).

**Figure 4.**
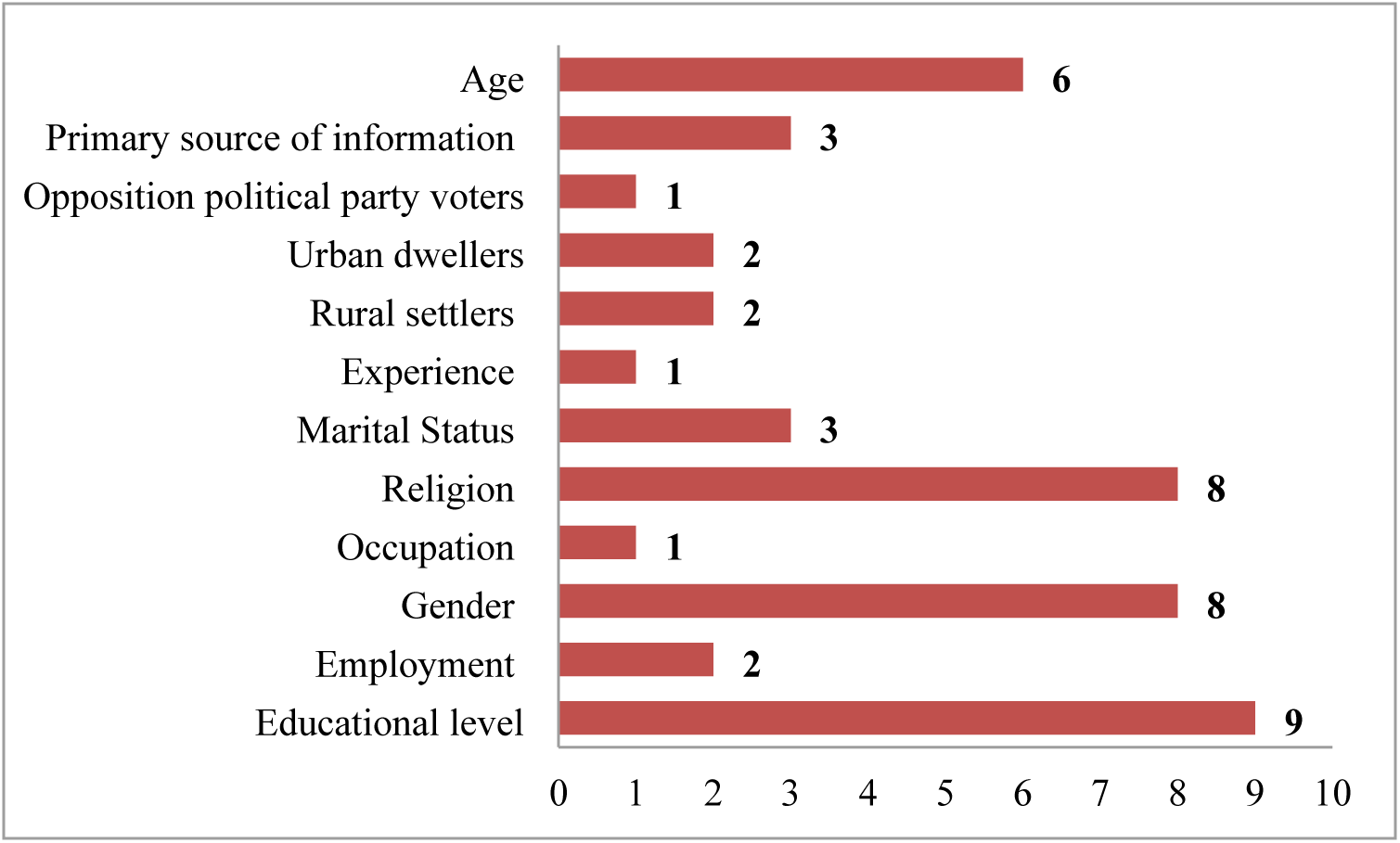
Evaluated demographic factors

**Table 7.** Demographics factors.

| **Demographics** | **Studies** | **Frequency** |
| --- | --- | --- |
| Educational level | [5], [8], [11], [13], [27], [34], [37], [49], [44] | 9 |
| Gender | [6], [8], [11], [30], [35], [37], [42], [44] | 8 |
| Religion | [8], [13], [15], [27], [34] [42], [44], [49] | 8 |
| Age | [9], [11], [15], [27], [42], [49] | 6 |
| Marital Status | [12], [37], [42] | 3 |
| Primary source of information about COVID-19 vaccine | [11], [13], [44] | 3 |
| Employment | [5], [37] | 2 |
| Rural settlers | [27], [37] | 2 |
| Urban dwellers | [27], [44] | 2 |
| Occupation | [7] | 1 |
| Experience | [15] | 1 |
| Opposition political party voters | [44] | 1 |

According to Backhaus [50], urban residency and current pregnancy were significantly and positively associated with refusing vaccination against COVID-19, whereas age, savings, and using contemporary contraceptives were significantly and negatively associated with refusing vaccination. Additional research by Hwang et al. [51] indicates that younger age, lack of religious affiliation, political conservatism, and lower family income are also substantially associated with vaccine hesitancy. For vaccine acceptance, [18] highlight that gender; age, education, and occupation are some of the socio-demographic variables associated with vaccine acceptance.

#### 3.4.4 What are the main study approaches utilized in examining the factors influencing the acceptance and hesitation of COVID-19 vaccine in Ghana?

The primary research methodology employed to investigate the factors influencing the acceptance and hesitancy of the COVID-19 vaccine in Ghana has predominantly been the quantitative approach, as indicated in Table 8. This was followed by the mixed-method approach.

**Table 8.** Study approaches used.

| Approach | Studies | Frequency |
| --- | --- | --- |
| Quantitative | [5-13], [15], [26-30], [33-37], [42], [44], [45], [47], [49] | 25 |
| Qualitative | [14], [31] | 2 |
| Mixed Method | [32], [43], [46], [48] | 4 |

Nevertheless, the review findings have brought attention to the insufficient utilization of the qualitative approach method in examining the factors that influence the acceptance and hesitancy towards the COVID-19 vaccine in Ghana (see Figure 5).

**Figure 5.**
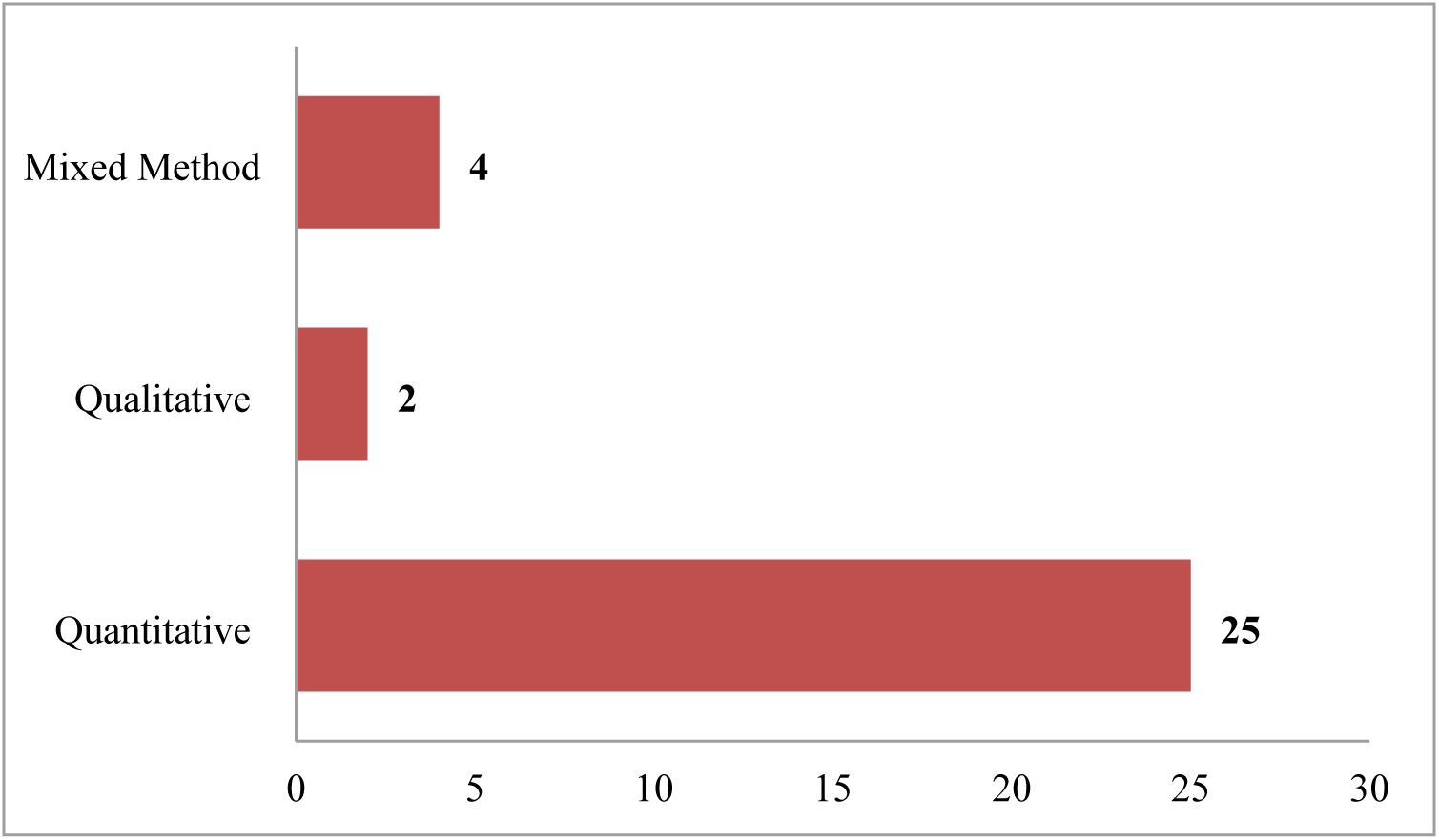
Study approaches

#### 3.4.5 What is the COVID-19 vaccine acceptance rate in Ghana?

The review that was conducted on the acceptance rate of the COVID-19 vaccine in Ghana shows the rate seen from September to October 2020 was highly promising among healthcare workers. However, a further study conducted from September to October 2020 on citizens above 18 years old in all 16 regions of Ghana shows a marginal decrease. The decline persisted until December 2020, as illustrated in Figure 6.

**Figure 6.**
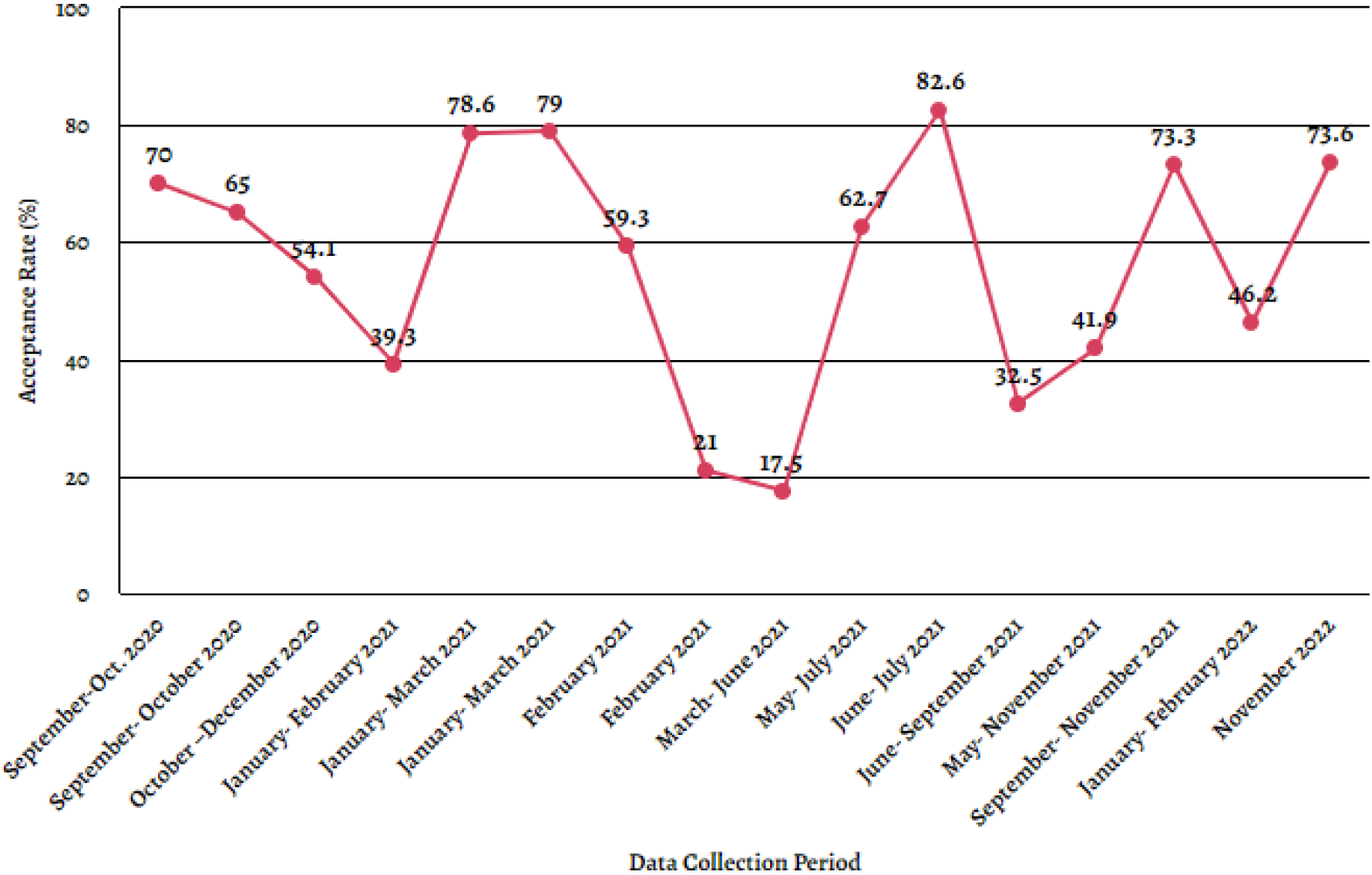
Acceptance rate (%)

During the first two months of the year 2021, there was a notable decrease in the rate of acceptance of COVID-19 vaccines among citizens in the 16 regions of Ghana. However, data collected from January to March 2021 show a high rate of acceptance of the COVID-19 vaccination (Kintampo North-Bono East Region). From February 2021 to June 2021, there was a sustained decline in the acceptance rate. It is imperative to recognize that between May and July 2021, there was a significant increase in the rate of acceptance of COVID-19 vaccines. Furthermore, it is noteworthy that the period spanning June to July 2021 had the highest reported level of acceptance of COVID-19 vaccines. However, there was a decrease in the COVID-19 vaccine acceptance rate in September 2021. In contrast, November had a slight increase in vaccine acceptance rate compared to September 2021. It is important to acknowledge that there was a decrease in the rate of acceptance of COVID-19 vaccines from January to February 2022. However, it is noteworthy that by November 2022, there had been a significant increase in the acceptance rate of COVID-19 vaccines in Ghana.

Table 9 illustrates that the main population group targeted for measuring the acceptance rate of COVID-19 vaccines in Ghana consisted predominantly of individuals aged 18 years and above, as well as healthcare professionals. It is noteworthy to mention that the population also encompassed registered radiographers, OPD attendants, government officials, community leaders, inhabitants in remote communities, as well as parents and guardians.

**Table 9.** Data Collection Period vs. Acceptance Rate.

| Selected Studies | Data Collection Period | Acceptance Rate (%) | Population Type | Area |
| --- | --- | --- | --- | --- |
| [8] | September-October 2020 | 70 | Health care workers | 16 Regions in Ghana |
| [12] | September- October 2020 | 65 | Citizens above 18 years | 16 Regions in Ghana |
| [36] | October –December 2020 | 54.1 | Adult Ghanaians | 16 Regions in Ghana |
| [30] | January- February 2021 | 39.3 | Health care workers | 16 Regions in Ghana |
| [29] | January- March 2021 | 78.6 | Health care workers | Kintampo North |
| [27] | January- March 2021 | 79 | Adult Ghanaians | 16 Regions in Ghana |
| [13] | February 2021 | 59.3 | Registered Radiographers | 16 Regions in Ghana |
| [11] | February 2021 | 21 | Adult Ghanaians | 16 Regions in Ghana |
| [37] | March- June 2021 | 17.5 | Citizens above 18 years | 16 Regions in Ghana |
| [7] | May- July 2021 | 62.7 | Adult Ghanaians | Dodowa |
| [9] | June- July 2021 | 82.6 | OPD attendants | Bono Region |
| [46] | June- September 2021 | 32.5 | Government Officials and community leaders | Volta Region |
| [5] | May- November 2021 | 41.9 | Residents in Rural Communities | Northern Region/ Ashanti Region/ Western North Region |
| [28] | September- November 2021 | 73.3 | Parents and Guardians | 16 Regions in Ghana |
| [26] | January- February 2022 | 46.2 | Citizens above 18 years | 16 Regions in Ghana |
| [15] | November 2022 | 73.6 | Health care workers | Bono Region |

The data summarized in Table 9 also provides additional information regarding the study of the COVID-19 vaccine acceptance rate in Ghana. The findings show that the majority of the reviewed studies explicitly concentrated on all 16 regions in the country. Additionally, certain locations within specific regions, such as Kintampo North, Dodowa, Bono Region, Volta Region, Northern Region, Ashanti Region, and Western North Region, were also included in the analysis.

## 4. Conclusion

This study presents a systematic review of the domain of COVID-19 vaccine acceptance and vaccine hesitancy in Ghana. It aims to investigate the factors that contribute to the acceptance of the COVID-19 vaccine as well as the contributing causes of vaccine hesitancy in the country. Furthermore, it examines the demographic factors commonly evaluated when determining the factors that influence the acceptance and hesitancy of the COVID-19 vaccine in Ghana. The study also explores the main study approaches utilized in examining these factors. Lastly, it provides an overview of the COVID-19 acceptance rate in Ghana. First, a total of 31 scholarly articles were identified, which were published throughout the timeframe of 2021 to April 2023. This identification process involved the implementation of a systematic approach, encompassing a number of methodical procedures and rigorous evaluations to ensure the quality of the selected papers. Following a thorough screening process to determine eligibility, we conducted a thorough analysis and synthesis of the collected data extracted from the chosen studies pertaining to various facets of COVID-19 vaccine research. These aspects encompassed the acceptance and hesitancy among individuals, the overall acceptance rate, the research approach employed in the respective studies, as well as the demographic characteristics considered. The relevant results are as follows:

i. There were a total of twenty-six (26) factors identified as influential in influencing the acceptance of the COVID-19 vaccine in Ghana. Among these factors, key determinants included a positive perception of the vaccine, confidence in its safety and efficacy, adequate knowledge about COVID-19, a favorable attitude towards vaccination, the desire to protect oneself and one’s family, gender, observing others receiving the vaccine, willingness to be vaccinated, and trust in the accuracy of COVID-19-related information.
ii. The main reasons for COVID-19 vaccine hesitancy in Ghana were twenty-eight (28) contributing causes, with the main ones being unfavorable vaccine side effects, mistrust in the vaccine, lack of confidence in its safety, fear, spiritual and religious beliefs, inadequate logistics, lack of information, uncertainty, conspiracy theories and worries about its impact on the Ghanaian race, doubt about the vaccine’s efficacy, and fertility concerns.
iii. Full lists of twelve (12) demographic characteristics were found as routinely assessed while examining the determinants of COVID-19 vaccine acceptance and hesitancy in Ghana. The most prevalent are educational level, gender, religion, age, and marital Status.
iv. Among the selected corpus of scholarly literature, comprising thirty-one (31) publications, a majority of twenty-five (25) studies employed a quantitative methodology to investigate the determinants of acceptance and hesitancy towards COVID-19 vaccine in the context of Ghana.
v. The acceptance rates that exhibited the greatest values were 82.6%, 79%, 78.6%, 73.6%, and 73.3%.

This review offers guidelines and recommendations for researchers, particularly those who are new in the field, regarding the subject of COVID-19 vaccine acceptability and hesitancy in Ghana. It explores the diverse factors that influence the acceptance of these vaccines as well as the factors that contribute to vaccine hesitancy. Additionally, it shows appropriate research methods to be employed when conducting studies in this area. Finally, conclusions are made regarding the demographic factors to consider, as these will aid in the accurate measurement of vaccine acceptance and hesitancy. In our future work, we intend to examine the rate of vaccine hesitancy within the Ghanaian setting, while also incorporating data from other West African countries in our systematic review.

## Data Availability

All relevant data are within the manuscript and its Supporting Information files.

## Author Contributions

**Conceptualization:** Godwin Banafo Akrong, Rosemond Akpene Hiadzi.

**Data curation:** Godwin Banafo Akrong, Rosemond Akpene Hiadzi, Antonia Bernadette Donkor, Daniel Kwasi Anafo.

**Formal analysis:** Godwin Banafo Akrong.

**Funding acquisition:** The author(s) received no specific funding for this work.

**Methodology:** Godwin Banafo Akrong, Rosemond Akpene Hiadzi, Antonia Bernadette Donkor, Daniel Kwasi Anafo.

**Project administration:** Godwin Banafo Akrong.

**Supervision:** Rosemond Akpene Hiadzi, Antonia Bernadette Donkor, Daniel Kwasi Anafo.

**Validation:** Godwin Banafo Akrong, Rosemond Akpene Hiadzi.

**Writing – original draft:** Godwin Banafo Akrong.

**Writing – review & editing:** Godwin Banafo Akrong, Rosemond Akpene Hiadzi, Antonia Bernadette Donkor, Daniel Kwasi Anafo.

## Notes

### Competing Interest Statement

The authors have declared no competing interest.

